# Decreased diastolic hydraulic forces incrementally associate with survival beyond conventional measures of diastolic dysfunction

**DOI:** 10.1101/2021.12.22.21268283

**Authors:** Dhnanjay Soundappan, Angus SY Fung, Daniel E Loewenstein, David Playford, Geoffrey Strange, Rebecca Kozor, James Otton, Martin Ugander

**Affiliations:** Kolling Institute, Royal North Shore Hospital, and University of Sydney, Sydney, Australia; St Vincent’s Clinical School, University of New South Wales, Sydney, Australia; Department of Clinical Physiology, Karolinska University Hospital, and Karolinska Institutet, Stockholm, Sweden; School of Medicine, University of Notre Dame, Fremantle, Australia; Faculty of Medicine and Health, University of Sydney, Sydney, Australia; Department of Cardiology, Liverpool Hospital, University of New South Wales, Liverpool, Australia

## Abstract

Decreased hydraulic forces during diastole contribute to reduced left ventricular (LV) filling and heart failure with preserved ejection fraction. However, their association with diastolic function and patient outcomes are unknown. The aim of this study was to determine the association between diastolic hydraulic forces, estimated by echocardiography as the atrioventricular area difference (AVAD), and both diastolic function and survival. Patients (n=5,176, median [interquartile range] 5.0 [5.0–5.0] years follow-up, 1,213 events) were selected from the National Echo Database Australia based on the presence of relevant transthoracic echocardiographic measures, LV ejection fraction (LVEF) ≥50%, heart rate 50-100 beats/minute, the absence of moderate or severe valvular disease, and no prior cardiac surgery. AVAD was calculated as the cross-sectional area difference between the LV and left atrium. LV diastolic dysfunction was graded according to 2016 guidelines. AVAD was weakly associated with E/e’, left atrial volume index, and LVEF (multivariable global R2=0.15, p<0.001), and not associated with e’ and peak tricuspid regurgitation velocity. Decreased AVAD was independently associated with poorer survival, and demonstrated improved model discrimination after adjustment for diastolic function grading (C-statistic [95% confidence interval] 0.644 [0.629–0.660] vs 0.606 [0.592–0.621], p<0.001) and E/e’ (0.649 [0.635–0.664] vs 0.634 [0.618–0.649], p<0.001), respectively. Therefore, decreased hydraulic forces, estimated by AVAD, are weakly associated with diastolic dysfunction and demonstrate an incremental prognostic association with survival beyond conventional measures used to grade diastolic dysfunction.

## Introduction

Diastolic dysfunction contributes to the development of elevated left ventricular filling pressures and subsequent heart failure with preserved ejection fraction (HFpEF). Consequently, the physiological mechanisms underlying diastolic function remain an area of ongoing research in an attempt to develop effective therapeutic interventions for patients with HFpEF. Diastolic function is a composite description of the interaction between the mechanisms driving LV filling and the passive tension opposing LV filling which are responsible for lengthening the cardiomyocytes, increasing the volume of the LV, and generating the atrioventricular pressure gradient ^1,2^. The term diastolic dysfunction can refer to mechanical abnormalities present during diastole which impair ventricular relaxation and filling ^3^.

Hydraulic forces have recently been identified as a mechanism contributing to LV diastolic function ^4^. Hydraulic force is calculated as the product of the pressure in a liquid, and the surface area with which that liquid is in contact, in accordance with Pascal’s Law ^5^. Since the blood pressure in both chambers of the left heart is nearly identical during diastole ^6^, the difference in cross-sectional short-axis area between the LV and left atrium (LA) provides the geometric basis for a net hydraulic force exuded on the atrioventricular plane in the apex-to-base direction (Figure 1). This difference is termed the atrioventricular area difference (AVAD), and can be used as a surrogate measure of net hydraulic force. Cardiac anatomy in healthy volunteers has already been shown to demonstrate a net hydraulic force directed towards the LA contributing to LV filling during most of diastole ^4^. This force is expected to contribute to the diastolic longitudinal motion of the atrioventricular plane, facilitating LV lengthening and accounting for between 10% and 60% of the peak driving force of LV filling in healthy subjects ^4^. Applying these findings to a clinical population, a recent study investigating changes to diastolic hydraulic forces in patients with pathological cardiac remodelling found that patients with HFpEF had a smaller AVAD compared to healthy controls throughout diastole, possibly limiting LV filling and contributing to the diastolic dysfunction ^7^.

**Figure 1.**
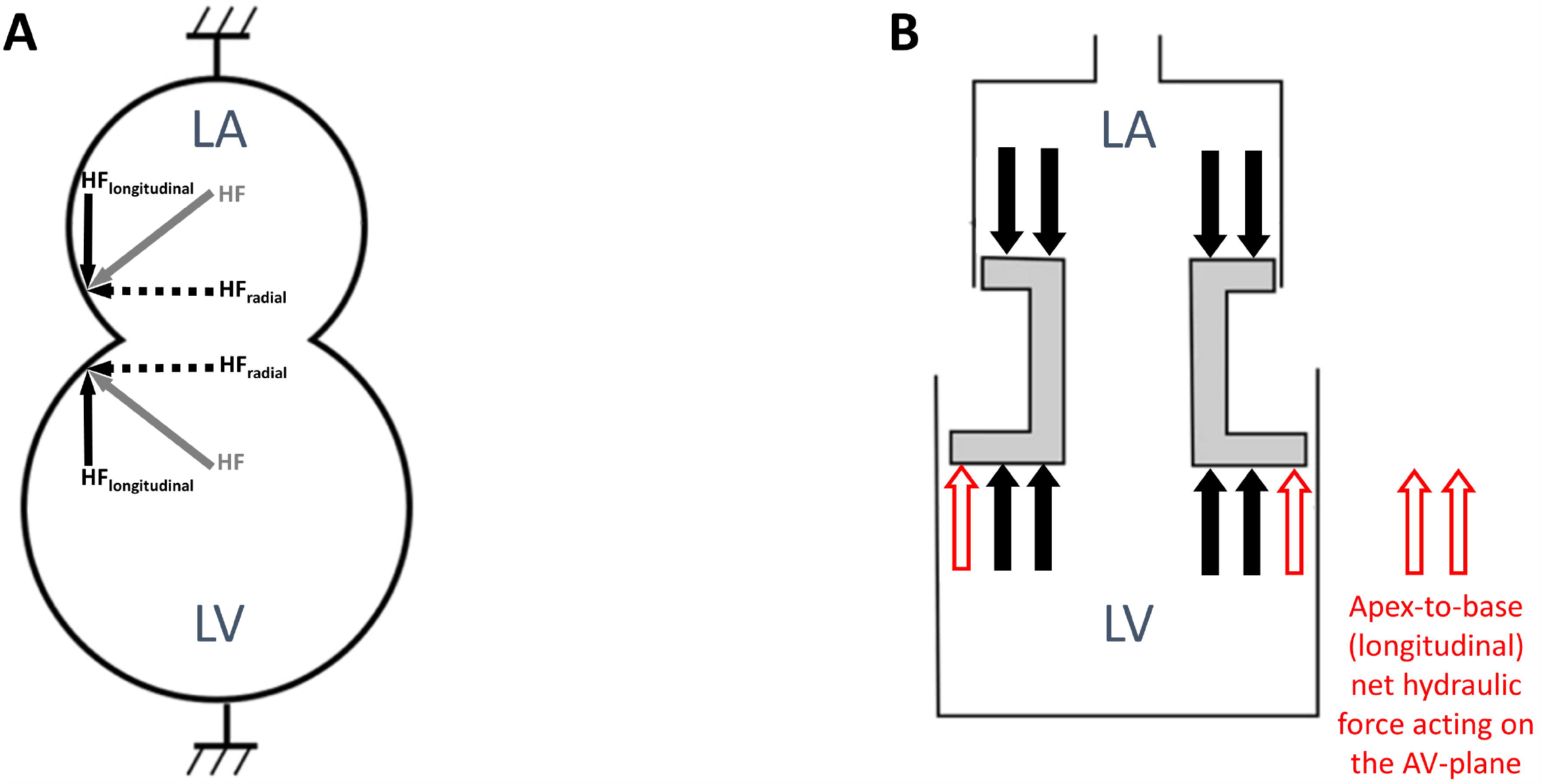
Schematic of hydraulic forces in the left atrium and ventricle. **(A)** During diastole, a hydraulic force (HF) is generated in the left atrium and ventricle perpendicular to the respective chamber walls, represented by the grey arrows in the left atrium and the left ventricle, respectively. These forces can be resolved into their longitudinal (HF_longitudinal_) and radial (HF_radial_) components as indicated by the solid black and dashed arrows, respectively. The radial component is counteracted by the pericardium and surrounding tissues whilst the longitudinal component represents the hydraulic force that contributes to the longitudinal motion of the atrioventricular plane during diastole. **(B)** Due to the larger surface area of the LV compared to the LA in a healthy individual, the LV generates a greater hydraulic force in the longitudinal direction, represented by the two additional red arrows. Abbreviations: HF = hydraulic force; HF_longitudinal_ = hydraulic force longitudinal component; HF_radial_ = hydraulic force radial component; LA = left atrium; LV = left ventricle. Figure adapted from ^4^.

Although hydraulic forces have been demonstrated to be a physiological mechanism contributing to diastolic filling in healthy and diseased states, their association with diastolic function and patient outcomes remains unknown. Furthermore, if a decrease in hydraulic force is independently associated with survival, this may be a potential therapeutic target in HFpEF. Specifically, established methods for LA reduction surgery may provide an opportunity to decrease LA size relative to LV size, in an attempt to aid the contribution of hydraulic forces to LV filling. Therefore, the aim of this study was to determine the association between diastolic hydraulic forces and both diastolic function and survival. We hypothesised that decreased diastolic hydraulic forces would be associated with diastolic dysfunction and poorer survival.

## Methods

### Study Design

The study population was derived using data from the National Echo Database Australia (NEDA). NEDA is an observational registry comprising retrospectively and prospectively collected digital transthoracic echocardiographic measurements from patients referred to laboratories across Australia since 1985, and is linked to health outcome data ^8^. The study complies with the Declaration of Helsinki, and has received approval from the lead ethics committee at the Royal Prince Alfred Hospital, Camperdown, Sydney, Australia (2019/ETH06989), and the human research ethics committees of all participating centers in Australia, respectively. The inclusion criteria for the study cohort were the presence of relevant echocardiographic measures, a LV ejection fraction (LVEF) greater than or equal to 50%, sinus rhythm, and a heart rate between 50 and 100 beats per minute. Echocardiographic measures of interest included LV end-diastolic diameter, LA end-systolic diameter, and at least two measures of diastolic function [E to septal e’ velocity ratio (E/e’), septal e’ velocity (e’ velocity), LA volume index (LAVI), or tricuspid regurgitation peak velocity]. These measures were selected as they are the recommended variables in the 2016 American Society of Echocardiography (ASE) and the European Association of Cardiovascular Imaging (EACVI) guidelines for evaluating LV diastolic function in patients ^9^. Exclusion criteria included moderate or severe valvular heart disease, and prosthetic valves. If multiple echocardiograms were available for a single patient, only the earliest recorded echocardiogram was included. The patient inclusion flowchart is described in Figure 2.

**Figure 2.**
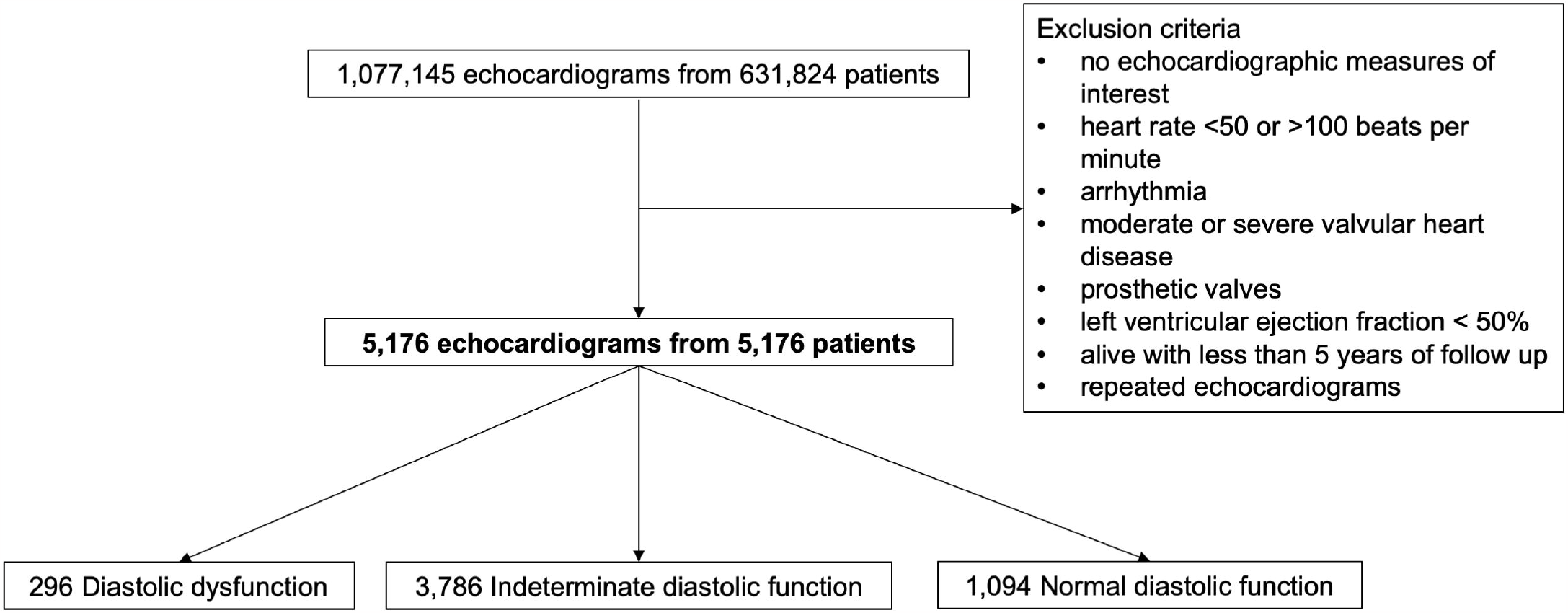
Patient inclusion flowchart. This flowchart describes the exclusion criteria of the study cohort.

### Image analysis

AVAD was calculated as the difference between LV and LA cross-sectional areas. LV and LA cross-sectional area were calculated by circular approximation using LV end-diastolic diameter and LA end-systolic diameter, respectively (Figure 3).

**Figure 3.**
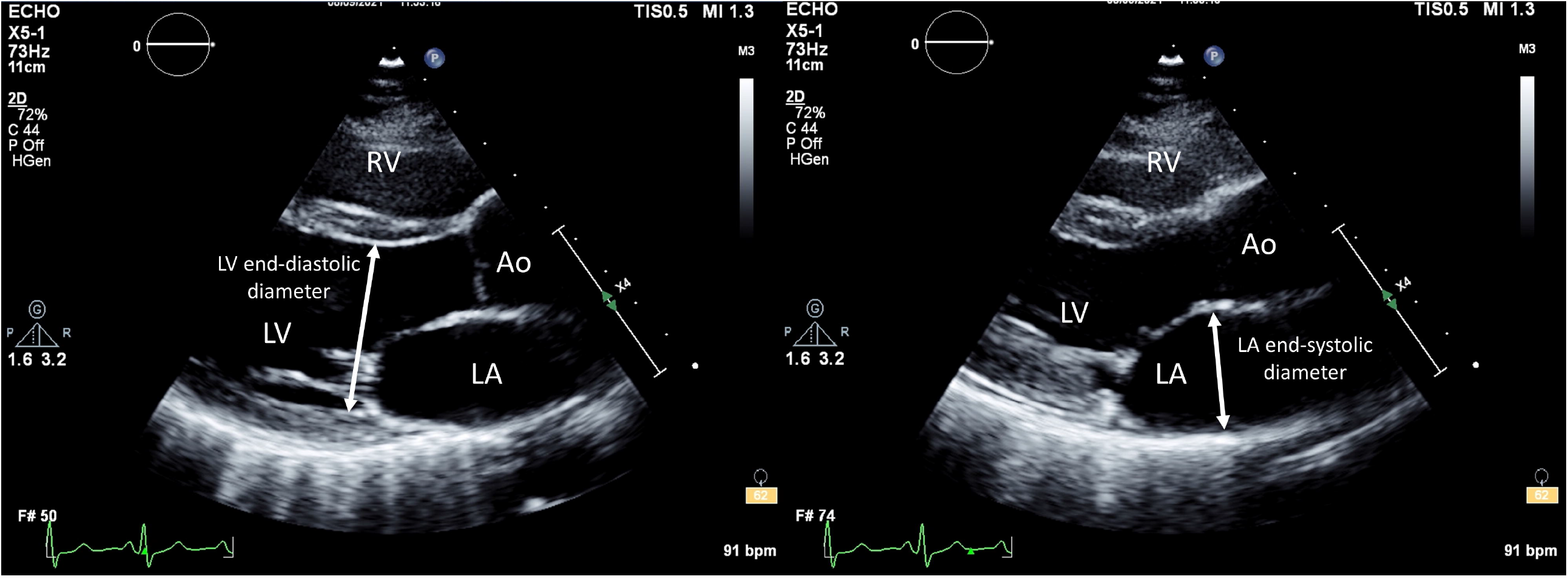
Measurement of atrioventricular area difference in transthoracic echocardiography. In routine echocardiography, atrioventricular area difference can be calculated as the difference between left ventricular end-diastolic short-axis cross-sectional area and left atrial end-systolic short-axis cross-sectional area. Short-axis cross-sectional area is calculated by circular approximation using left ventricular end-diastolic diameter and left atrial end-systolic diameter measured in a parasternal view. The atrial short-axis and ventricular short-axis areas represent the surface areas which contribute to the generation of hydraulic forces which contribute to the diastolic longitudinal motion of the atrioventricular plane. Abbreviations: Ao = aorta; LA = left atrium; LV = left ventricle; RV = right ventricle.

For all measures, the designation of end diastole and end systole refers to LV systole and diastole. Using any available measures of diastolic function, patients were graded using the 2016 ASE/EACVI guidelines ^9^. Specifically, patients were graded as having normal diastolic function if less than half of the available parameters met cut off values, indeterminate diastolic function if exactly half met cut off values, and diastolic dysfunction if more than half met cut off values.

### Statistical Analysis

All statistical analyses were conducted using programming language, R (R Core Team, R Foundation for Statistical Computing, Vienna, Austria). The characteristics of the study cohort were calculated and reported as the median [interquartile range]. Differences in measured variables between patients were assessed using Wilcoxon rank-sum test.

Univariable and multivariable linear regression were used to determine the relationship between AVAD and parameters of diastolic function in patient subgroups based on the availability of diastolic function measures. R^2^ values were used to represent the proportion of the variation in AVAD explained by the variables, whilst standardised beta coefficients were used to compare the strength of association with AVAD between variables. Survival analysis was conducted to examine the association between selected variables and time to death using Kaplan-Meier curves and Cox proportional hazards models. The end-points used were 5-year all-cause mortality and 5-year cardiovascular mortality. Univariable Cox regression models were used to evaluate the association between survival and AVAD, E/e’, and diastolic function grading, in the overall population and both sexes. Multivariable Cox regression models were used to evaluate the association between AVAD and survival, adjusting for conventional parameters of diastolic function. Finally, multivariable analyses were repeated in patient subgroups stratified by clinically relevant LVEF ranges. The proportional hazards assumption was confirmed by visual inspection of the Schoenfeld residuals, and by demonstrating a non-significant relationship between the residuals and time. Wald’s chi-square values were reported to compare the strength of association of different variables within regression models. Hazard ratios for continuous variables were scaled by standard deviation to allow for comparison between models, whilst hazard ratios for diastolic function grading were reported using normal diastolic function as a reference level. The goodness of fit of univariable and multivariable models was compared using the concordance (C) statistic, and differences in C-statistics were evaluated using a one-shot nonparametric approach independent of any overlap between the 95% confidence intervals of the two C-statistics being compared ^10^. A p-value less than 0.05 was considered statistically significant.

## Results

### Study population

The population considered for this study consisted of 5,176 patients, with cohort characteristics presented in Table 1. Patients were followed up for 5.0 [5.0–5.0] years, with 1,213 deaths occurring over this period. All measures of diastolic function were consistent with better diastolic function in patients with an AVAD above the median, however not by a clinically meaningful magnitude (p<0.001 for all).

**Table 1.**
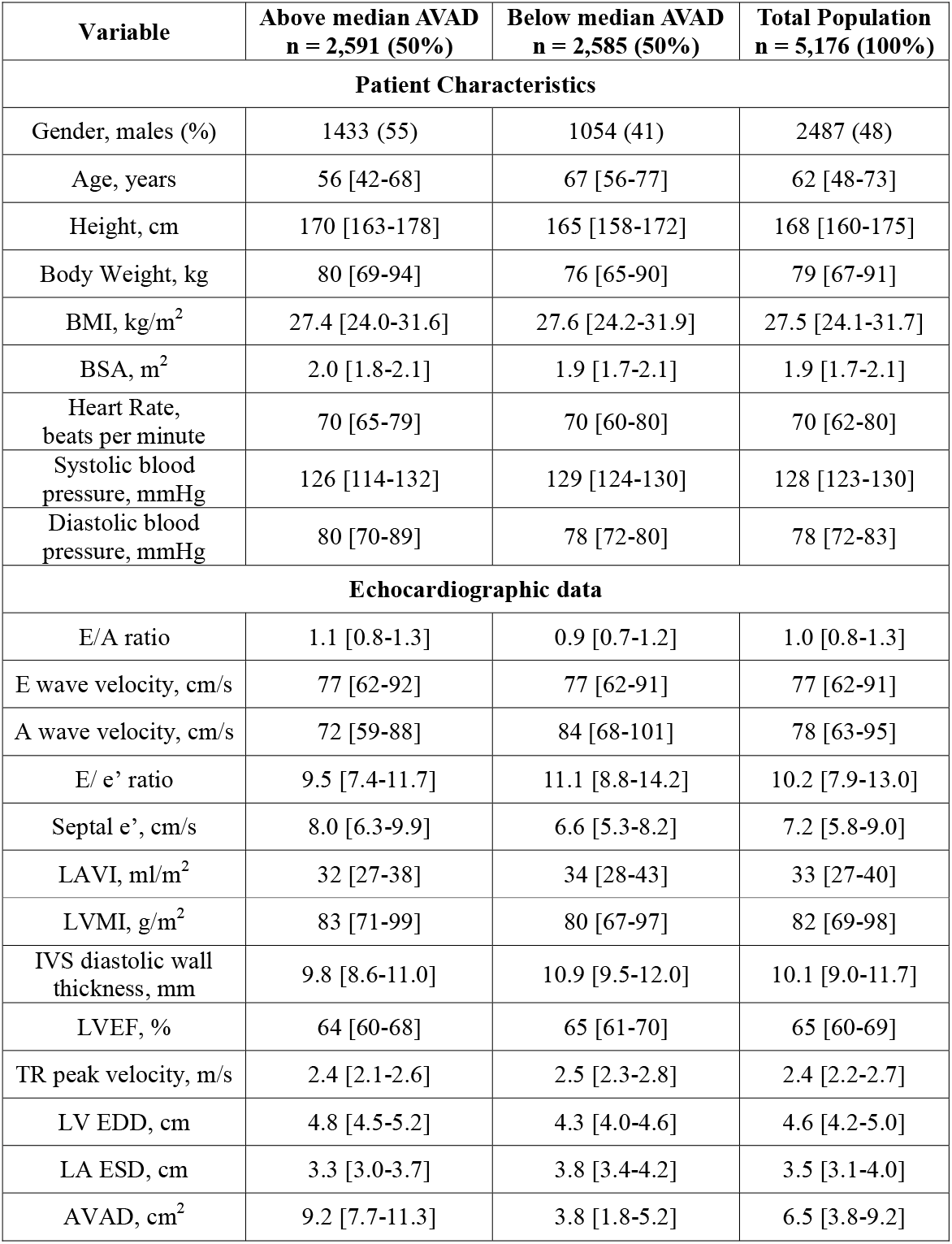
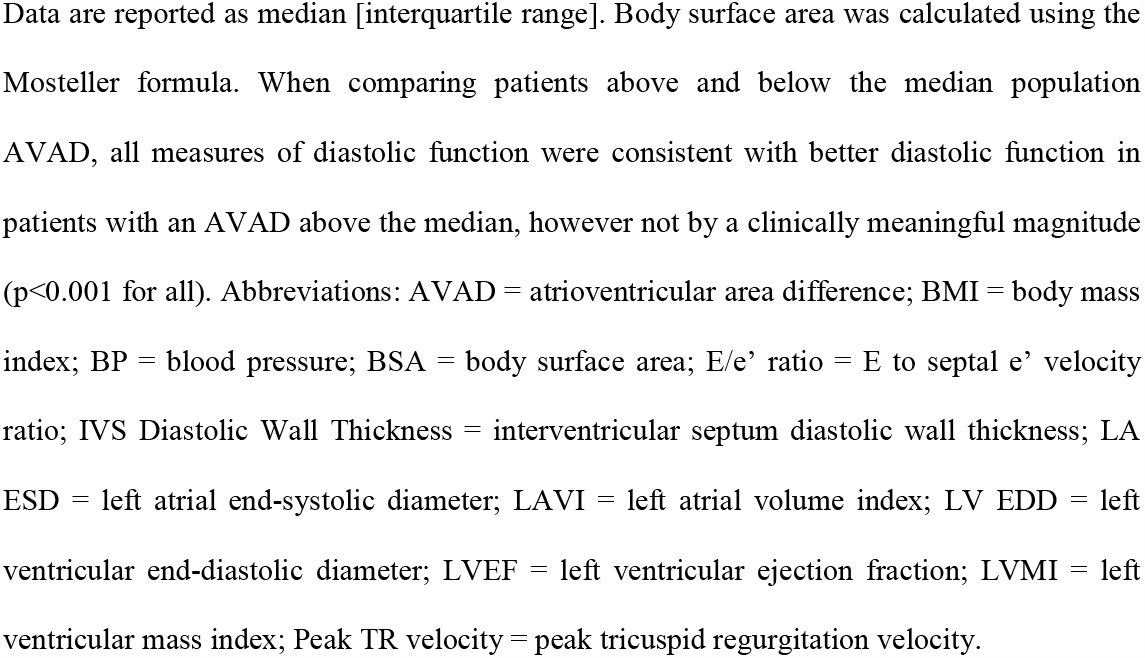
Baseline characteristics of study cohort.

| Variable | Above median AVAD<br>n = 2,591 (50%) | Below median AVAD<br>n = 2,585 (50%) | Total Population<br>n = 5,176 (100%) |
| --- | --- | --- | --- |
| **Patient Characteristics** |  |  |  |
| Gender, males (%) | 1433 (55) | 1054 (41) | 2487 (48) |
| Age, years | 56 [42-68] | 67 [56-77] | 62 [48-73] |
| Height, cm | 170 [163-178] | 165 [158-172] | 168 [160-175] |
| Body Weight, kg | 80 [69-94] | 76 [65-90] | 79 [67-91] |
| BMI, kg/m<sup>2</sup> | 27.4 [24.0-31.6] | 27.6 [24.2-31.9] | 27.5 [24.1-31.7] |
| BSA, m<sup>2</sup> | 2.0 [1.8-2.1] | 1.9 [1.7-2.1] | 1.9 [1.7-2.1] |
| Heart Rate,<br>beats per minute | 70 [65-79] | 70 [60-80] | 70 [62-80] |
| Systolic blood<br>pressure, mmHg | 126 [114-132] | 129 [124-130] | 128 [123-130] |
| Diastolic blood<br>pressure, mmHg | 80 [70-89] | 78 [72-80] | 78 [72-83] |
| **Echocardiographic data** |  |  |  |
| E/A ratio | 1.1 [0.8-1.3] | 0.9 [0.7-1.2] | 1.0 [0.8-1.3] |
| E wave velocity, cm/s | 77 [62-92] | 77 [62-91] | 77 [62-91] |
| A wave velocity, cm/s | 72 [59-88] | 84 [68-101] | 78 [63-95] |
| E/ e' ratio | 9.5 [7.4-11.7] | 11.1 [8.8-14.2] | 10.2 [7.9-13.0] |
| Septal e', cm/s | 8.0 [6.3-9.9] | 6.6 [5.3-8.2] | 7.2 [5.8-9.0] |
| LAVI, ml/m<sup>2</sup> | 32 [27-38] | 34 [28-43] | 33 [27-40] |
| LVMI, g/m<sup>2</sup> | 83 [71-99] | 80 [67-97] | 82 [69-98] |
| IVS diastolic wall<br>thickness, mm | 9.8 [8.6-11.0] | 10.9 [9.5-12.0] | 10.1 [9.0-11.7] |
| LVEF, % | 64 [60-68] | 65 [61-70] | 65 [60-69] |
| TR peak velocity, m/s | 2.4 [2.1-2.6] | 2.5 [2.3-2.8] | 2.4 [2.2-2.7] |
| LV EDD, cm | 4.8 [4.5-5.2] | 4.3 [4.0-4.6] | 4.6 [4.2-5.0] |
| LA ESD, cm | 3.3 [3.0-3.7] | 3.8 [3.4-4.2] | 3.5 [3.1-4.0] |
| AVAD, cm<sup>2</sup> | 9.2 [7.7-11.3] | 3.8 [1.8-5.2] | 6.5 [3.8-9.2] |

Data are reported as median [interquartile range]. Body surface area was calculated using the Mosteller formula. When comparing patients above and below the median population AVAD, all measures of diastolic function were consistent with better diastolic function in patients with an AVAD above the median, however not by a clinically meaningful magnitude ( $p < 0.001$ for all). Abbreviations: AVAD = atrioventricular area difference; BMI = body mass index; BP = blood pressure; BSA = body surface area; E/e' ratio = E to septal e' velocity ratio; IVS Diastolic Wall Thickness = interventricular septum diastolic wall thickness; LA ESD = left atrial end-systolic diameter; LAVI = left atrial volume index; LV EDD = left ventricular end-diastolic diameter; LVEF = left ventricular ejection fraction; LVMI = left ventricular mass index; Peak TR velocity = peak tricuspid regurgitation velocity.

### Atrioventricular area difference and diastolic function

AVAD was univariably associated with E/e’, e’ velocity, LAVI, peak tricuspid regurgitation velocity and LVEF. In multivariable linear regression, only E/e’, LAVI and LVEF remained associated with AVAD, with an overall weak association (global adjusted R^2^=0.15, p<0.001, Table 2).

**Table 2.** Transthoracic echocardiographic measures of diastolic function and their association with atrioventricular area difference.

|  | **Univariable** |  | **Multivariable**<br>**(319 patients)** |  |  |  |
| --- | --- | --- | --- | --- | --- | --- |
| **Measure** | **R<sup>2</sup>** | **p value** | **Standardised $\beta$** | **p value** | **Global Adjusted R<sup>2</sup>** | **p value** |
| E/e' ratio<br>(5,176 patients) | 0.06 | <0.001 | -0.31 | <0.001 | 0.15 | <0.001 |
| e' velocity, cm/s<br>(4,747 patients) | 0.05 | <0.001 | -0.05 | 0.41 |  |  |
| LA volume index, cm<sup>3</sup>/m<sup>2</sup><br>(957 patients) | 0.03 | <0.001 | -0.12 | 0.04 |  |  |
| TR peak velocity, cm/s<br>(2,731 patients) | 0.05 | <0.001 | -0.02 | 0.71 |  |  |
| LVEF, %<br>(5,176 patients) | 0.03 | <0.001 | -0.19 | <0.001 |  |  |

R<sup>2</sup> values represent the proportion of the variation in AVAD explained by the variables, whilst standardised beta coefficients express the strength of the effect of each independent variable on AVAD. The number of patients included in each respective analysis is included in brackets.
Abbreviations: AVAD = atrioventricular area difference; e' velocity = septal e' velocity; E/e' = E to septal e' velocity ratio; LAVI = left atrial volume index; LVEF = left ventricular ejection fraction; Peak TR velocity = peak tricuspid regurgitation velocity. TR peak velocity = tricuspid regurgitation peak velocity; LVEF = left ventricular ejection fraction, LA ESD = left atrial end systolic diameter, LV EDD = left ventricular end diastolic diameter.

### Prognostic value of atrioventricular area difference

The results of univariable and multivariable Cox regression analyses using 5-year all-cause mortality as the outcome measure are summarised in Tables 3 and 4. In univariable Cox regression, AVAD was positively associated with survival (hazard ratio (HR) [95% confidence interval (CI)] 1.33 [1.26–1.40] per SD decrement, p<0.001, Table 3) and E/e’ was negatively associated with survival (HR [95% CI] 1.49 [1.43–1.56] per SD increment, p<0.001, Table 4). Both diastolic dysfunction and indeterminate diastolic function were associated with poorer survival compared to normal diastolic function (HR [95% CI] 2.99 [2.63–3.39], p<0.001 for diastolic dysfunction). These results were replicated in sex-disaggregated analysis (data not shown). The Kaplan-Meier survival curves for these univariable analyses are represented in Figure 4. In multivariable analyses, AVAD demonstrated improved model discrimination when included with diastolic function grading (C-statistic [95% CI] 0.644 [0.629–0.660] vs 0.606 [0.592–0.621], p<0.001, Table 3) and E/e’ (C-statistic [95% CI] 0.649 [0.635–0.664] vs 0.634 [0.618–0.649], p<0.001, Table 4) respectively. These results were unchanged when cardiovascular mortality was used as the outcome measure (Supplemental Tables 1 and 2). Additionally, a similar trend in results was observed when patients were stratified into subgroups of 50% ≤ LVEF < 60%, 60% ≤ LVEF < 75% and 75% ≤ LVEF (Supplemental Tables 3 and 4).

**Table 3.** Atrioventricular area difference and diastolic function grading as predictors of 5-year all-cause mortality.

| LVEF ≥ 50%<br>n = 5176, 1213 events<br>5.0 [5.0–5.0] yrs follow up | Univariable Model | | | | Multivariable Model | | | |
| --- | --- | --- | --- | --- | --- | --- | --- | --- |
| Variable | Chi-Square | HR [95% CI] | p value | C-statistic [95% CI] | Chi-Square | HR [95% CI] | p value | C-statistic [95% CI] |
| Diastolic Dysfunction | 288 | 2.99<br>[2.63 – 3.39] | <0.001 | 0.606<br>[0.592 – 0.621] | 208 | 2.64<br>[2.32 – 3.02] | <0.001 | 0.644<br>[0.629 – 0.660] |
| Indeterminate Diastolic Function | 22 | 1.48<br>[1.26 – 1.74] | <0.001 |  | 17 | 1.42<br>[1.20 – 1.67] | <0.001 |  |
| AVAD | 111 | 1.33<br>[1.26 – 1.40] | <0.001 | 0.588<br>[0.573 – 0.604] | 41 | 1.20<br>[1.14 – 1.27] | <0.001 |  |

Cox regression was used to evaluate AVAD and diastolic function grading as predictors of 5-year all-cause mortality. Diastolic function grading was determined using the 2016 ASE/EACVI guidelines. Wald's chi-square values were used to compare the strength of association of different variables within regression models. The hazard ratios for diastolic dysfunction and indeterminate diastolic function are reported with normal diastolic function as a reference level. The hazard ratio for AVAD is scaled by standard deviation decrement. The C-statistic of univariable and multivariable models was compared to evaluate differences in model discrimination. Abbreviations: ASE = American Society of Echocardiography; AVAD = atrioventricular area difference; CI = confidence interval; EACVI = European Association of Cardiovascular Imaging; HR = hazard ratio; LVEF = left ventricular ejection fraction.

**Table 4.** Atrioventricular area difference and E/e’ as predictors of 5-year all-cause mortality.

| LVEF ≥ 50%<br>n = 5176, 1213 events<br>5.0 [5.0–5.0] yrs follow up | Univariable Model |  |  |  | Multivariable Model |  |  |  |
| --- | --- | --- | --- | --- | --- | --- | --- | --- |
| Variable | Chi-Square | HR [95% CI] | p value | C-statistic [95% CI] | Chi-Square | HR [95% CI] | p value | C-statistic [95% CI] |
| E/e' ratio | 319 | 1.49<br>[1.43 – 1.56] | <0.001 | 0.634<br>[0.618 – 0.649] | 225 | 1.43<br>[1.36 – 1.49] | <0.001 | 0.649<br>[0.635 – 0.664] |
| AVAD | 111 | 1.33<br>[1.26 – 1.40] | <0.001 | 0.588<br>[0.573 – 0.604] | 42 | 1.21<br>[1.14 – 1.28] | <0.001 |  |

Cox regression was used to evaluate AVAD and E/e' as predictors of 5-year all-cause mortality. Wald's chi-square values were used to compare the strength of association of different variables within regression models. The hazard ratio for E/e' ratio is scaled by standard deviation increment, and AVAD by standard deviation decrement. The C-statistic of univariable and multivariable models was compared to evaluate differences in model discrimination. Abbreviations: AVAD = atrioventricular area difference; CI = confidence interval; E/e' = E to septal e' velocity ratio; HR = hazard ratio; LVEF = left ventricular ejection fraction.

**Figure 4.**
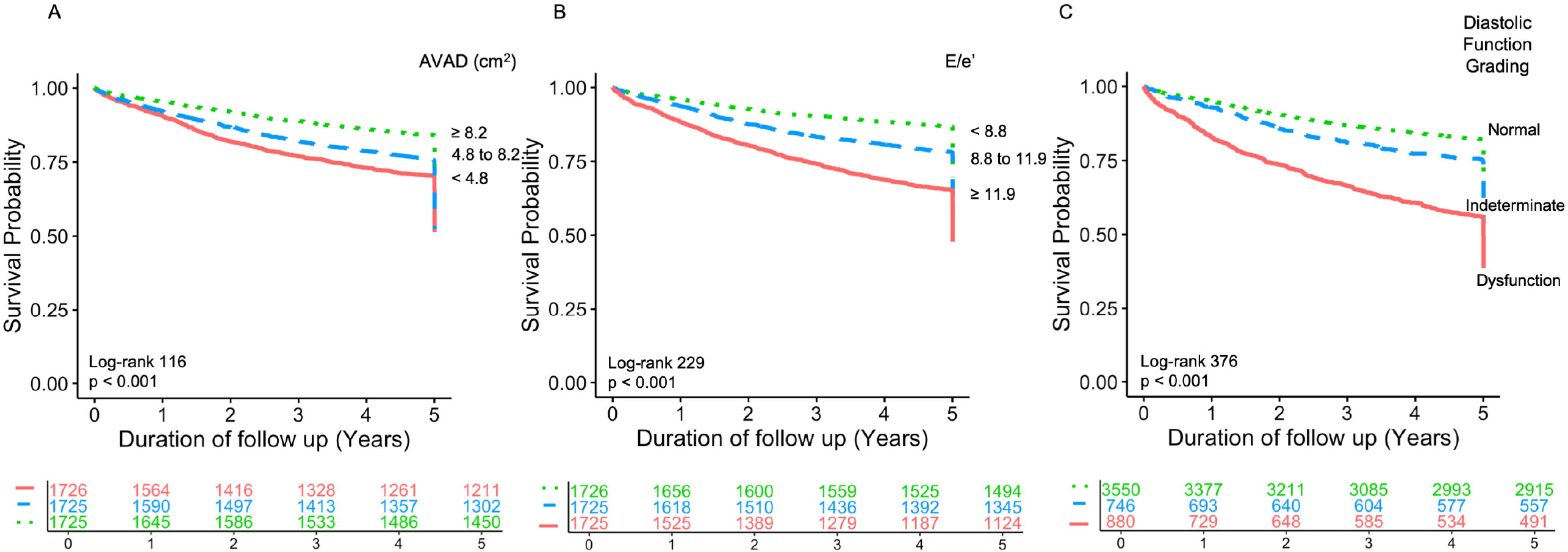
Survival curves for atrioventricular area difference, E/e’ ratio and diastolic function grading. Kaplan-Meier survival curves for (A) AVAD, (B) E/e’ and (C) diastolic function grading. Each curve is stratified by tertiles, with the respective risk table below each panel. Abbreviations: AVAD = atrioventricular area difference.

## Discussion

The main finding of this study is that decreased diastolic hydraulic forces, estimated by AVAD, are associated with poor survival. This association between diastolic hydraulic forces and survival is independent of, and incremental to, current measures of diastolic function. Taken together, this suggests that further investigation evaluating increased LA size relative to LV size as a potential therapeutic target in HFpEF is justified.

Diastolic hydraulic forces provide a macroscopic explanation to a part of the physiology behind diastolic function, incorporating the effects of blood pressure and cardiac geometry. The contribution of diastolic hydraulic forces to LV filling in health has been found to be of comparable magnitude to other diastolic mechanisms such as active relaxation and restoring forces ^4^. Meanwhile, conventional parameters recommended by the ASE/EACVI to grade diastolic function account for the contribution of LV early diastolic recoil, LV relaxation, LV filling pressure, and LA pressure ^11^. The recommended algorithm from these guidelines has been previously associated with increased cardiovascular-related and all-cause mortality ^12^. The current study shows that AVAD provides prognostic value beyond these measures to predict survival in a clinical population. When compared directly with E/e’, the echocardiographic parameter most closely related to LV filling pressure ^13,14^, AVAD retained an independent association with survival, albeit with a somewhat weaker strength of association. These results, including the finding of merely a weak association between AVAD and conventional diastolic measures, indicate that AVAD provides novel information when assessing diastolic function and prognosis in patients.

### Method for calculating AVAD

Our study calculated AVAD as the difference between approximated LV end-diastolic and LA end-systolic short-axis cross-sectional areas, respectively. Short-axis cross-sectional area was measured as it represents the two-dimensional surface area upon which hydraulic forces in the base-apex direction act on the mitral annulus during diastole. By comparison, while the third dimension of chamber length in the base-apex direction is essential for quantifying chamber volume, it does not influence the hydraulic forces in the base-apex direction that are acting upon the mitral annulus. Routine echocardiographic measurements of the LV are performed at end systole and end diastole, while the dimensions of the LA are measured only at end systole. However, the contribution of AVAD to the net diastolic hydraulic force is most accurately calculated during mid-diastasis when LA and LV pressures equalise ^4^.

Whilst there is no meaningful magnitude of change in LV cross-sectional area between mid-diastasis and end diastole, LA cross-sectional area is greater at end systole than at mid diastasis ^4^. Specifically, in a study of healthy volunteers, LA short-axis area increased from 14cm^2^ to 19cm^2^ between mid-diastasis and end systole ^4^. This change in area is related to the movement of the mitral annular plane, or mitral annular plane systolic excursion (MAPSE) ^15,16^. Specifically, as the mitral annular plane is displaced towards the apex during systole, LA cross-sectional area increases. To account for the potential influence of MAPSE on our measurement of AVAD, we repeated our statistical analyses in patients stratified by subgroups of LVEF above 50%, and observed a similar trend in results. It is well established that MAPSE is positively associated with LVEF ^17-20^. Therefore, it can be inferred that patients with a similar LVEF also have an adequately similar MAPSE and the movement of the mitral annular plane does not considerably affect the differences in AVAD between these patients. This is supported by the negligibly weak association found between AVAD and LVEF within each subgroup of LVEF above 50% (R^2^=0.01-0.03, p<0.001, Supplemental Table 7). As such, when comparing patients with similar LVEF, AVAD measured using LV end-diastolic diameter and LA end-systolic diameter can be used as a reasonable surrogate for diastolic hydraulic forces that ideally would be based on AVAD measured during mid-diastasis.

### The relative geometry of the left ventricle and left atrium

Cardiac geometry is known to play an integral role in cardiovascular health. Changes to the structure of the LV and LA, either independently or simultaneously, have been demonstrated to impair function and contribute to cardiovascular disease and outcomes ^21-27^. Whilst LA and LV measures are often studied separately, it has been suggested that a single parameter accounting for the complex interplay between LA and LV physiology may be a more clinically useful predictor of cardiovascular disease and survival ^28^. Extending on these ideas, it has recently been demonstrated that a left atrioventricular coupling index (LACI) was a strong predictor of cardiovascular events and death, with improved discrimination and reclassification power when compared to individual LA and LV measures ^28^. LACI was calculated by dividing LA end-diastolic volume by LV end-diastolic volume, with the authors acknowledging that LA and LV function and pressure are most closely related during diastole. Considering that cardiac chamber volume is dependent on mean cross-sectional area, the findings using LACI represent an approximate surrogate measure of diastolic hydraulic forces, albeit without specifically measuring the difference in LV and LA short-axis cross-sectional areas.

### The potential of left atrial reduction surgery

The independent prognostic value of AVAD indicates that increased LA size relative to LV size may be a new therapeutic target in patients with HFpEF. In HFpEF, elevated LV filling pressures are propagated to the LA, resulting in remodelling and dilatation ^22,27,29^. This may result in the LA cross-sectional area becoming equal to or larger than the LV cross-sectional area, orienting the net diastolic hydraulic force towards the apex of the heart and opposing LV filling ^7^. Decreasing LA size in these patients may potentially restore the contribution of diastolic hydraulic forces to LV filling, possibly improving both diastolic function and survival. LA reduction surgery is already an established procedure ^30-34^ primarily indicated alongside other surgical procedures to treat an enlarged LA or chronic atrial fibrillation in patients with mitral valve disease ^31^. Several studies have highlighted the benefits of LA reduction surgery, most commonly resulting in increased restoration and maintenance of sinus rhythm, with no significant increases in mortality or post-operative complications ^35-41^. The mechanism for these improvements is not clearly understood. However, it is thought to be related to electrophysiological remodelling and increased wall stress ^42,43^. We hypothesise that these benefits may also be attributed to the subsequent improvement in net diastolic hydraulic force and diastolic function. Diastolic dysfunction is thought to contribute to increased atrial afterload, stretching and wall stress, ultimately increasing the risk of atrial fibrillation ^44,45^. Considering the importance of cardiac geometry and diastolic function in HFpEF as well, the current results support the notion that LA reduction surgery concomitant with otherwise indicated open-heart surgery is a treatment option that merits prospective evaluation with regards to improving symptoms and outcomes in carefully selected patients with HFpEF.

### Limitations

As it currently stands, NEDA does not contain clinical data relating to patient comorbidities, cardiovascular disease risk factors, and pharmacotherapy. This information may impact results, as these clinical details affect both survival and diastolic function, especially since the NEDA population primarily consists of patients referred for echocardiography due to known or suspected cardiovascular disease. Nevertheless, echocardiographic data paired with survival from NEDA has been previously used to draw important prognostic information on pulmonary hypertension, aortic stenosis, and diastolic dysfunction, reflecting the robustness of using mortality end-points despite a lack of clinical data ^12,46-49^. Additionally, our methodology may have been limited by measurement variability inherent to the assumption of circular cross-sectional areas and the variability in selecting the locations of measuring LV and LA diameters. However, our ability to consistently demonstrate clear prognostic value using clinically acquired data despite this variability indicates the robustness of these prognostic relationships. Finally, it is important to consider inter-observer and inter-site repeatability, since NEDA data is collected from more than 20 echocardiography laboratories across Australia. This is accounted for by stringent NEDA protocols implemented during data acquisition and processing. Inspection of descriptive statistics of the included measures (data not shown) also did not indicate any reason for concern regarding systematic variations in the data. In light of the findings and limitations of the current study, future studies should look to further validate the prognostic value of AVAD, while overcoming the limitations of the current study by ideally measuring AVAD in mid diastole (diastasis), and including relevant clinical characteristics.

## Conclusions

A decrease in diastolic hydraulic forces, estimated by AVAD, is weakly associated with diastolic dysfunction, and independently associated with poorer survival in a clinical population referred for echocardiography. Increased LA size relative to the LV may impair the longitudinal motion of the atrioventricular plane during diastole, impairing LV filling and contributing to a poorer prognosis for patients with cardiovascular disease.

## Data availability

The data that support the findings of this study are available from the National Echo Database Australia but restrictions apply to the availability of these data, which were used under license for the current study, and so are not publicly available.

## Data Availability

All data produced in the present work are contained in the manuscript (unless otherwise specified). Supplementary data in the present study is available upon reasonable request to the authors.

## Acknowledgements

The authors acknowledge and thank Professor Håkan Arheden, Lund University, Sweden, for valuable discussions and input.

## Author Contributions

D.S., J.O., and M.U. were involved in analysis and interpretation of data, and drafting of the manuscript. All authors were involved in study design and have revised and approved the final manuscript.

## Additional Information

DP has received modest honorarium from Alerte Echo IQ as well as consultancy fees from NEDA. All other authors declare no competing interests.

