## Supplemental Data for "Decreased diastolic hydraulic forces incrementally associate with survival beyond conventional measures of diastolic dysfunction"

**Supplemental Table 1. Atrioventricular area difference and diastolic function grading as predictors of 5-year cardiovascular mortality.**

| LVEF ≥ 50% n = 5176, 173 events  5.0 [5.0–5.0] yrs follow up | **Univariable Model** | | | | **Multivariable Model** | | | |
| --- | --- | --- | --- | --- | --- | --- | --- | --- |
| **Variable** | Chi-Square | HR [95% CI] | p value | C-statistic  [95% CI] | Chi-Square | HR [95% CI] | p value | C-statistic  [95% CI] |
| Diastolic Dysfunction | 111 | 5.80 [4.18 – 8.03] | <0.001 | 0.687  [0.648 – 0.726] | 84 | 4.94 [3.51 – 6.95] | <0.001 | 0.718  [0.680 – 0.757] |
| Indeterminate Diastolic Function | 10 | 2.02 [1.29 – 3.17] | 0.002 |  | 8 | 1.91 [1.22 – 2.99] | 0.005 |  |
| AVAD | 37 | 1.51 [1.32 – 1.73] | <0.001 | 0.620  [0.580 – 0.661] | 10 | 1.27 [1.10 – 1.46] | 0.001 |  |

Univariable and multivariable cox regression was used to evaluate AVAD and diastolic function grading as predictors of 5-year cardiovascular mortality. Diastolic function grading was determined using the 2016 ASE/EACVI guidelines. Wald’s chi-square values were used to compare the strength of association of different variables within regression models. The hazard ratios for diastolic dysfunction and indeterminate diastolic function are reported with normal diastolic function as a reference level. The hazard ratio for AVAD is scaled by standard deviation decrement. The C-statistic of univariable and multivariable models was compared to evaluate differences in model discrimination. Abbreviations: ASE = American Society of Echocardiography; AVAD = atrioventricular area difference; CI = confidence interval; EACVI = European Association of Cardiovascular Imaging; HR = hazard ratio; LVEF = left ventricular ejection fraction.

**Supplemental Table 2. Atrioventricular area difference and E/e’ as predictors of 5-year cardiovascular mortality.**

| LVEF ≥ 50% n = 5176, 1213 events  5.0 [5.0–5.0] yrs follow up | **Univariable Model** | | | | **Multivariable Model** | | | |
| --- | --- | --- | --- | --- | --- | --- | --- | --- |
| **Variable** | Chi-Square | HR [95% CI] | p value | C-statistic  [95% CI] | Chi-Square | HR [95% CI] | p value | C-statistic  [95% CI] |
| E/e’ ratio | 86 | 1.64 [1.48 – 1.82] | <0.001 | 0.675  [0.634 – 0.716] | 56 | 1.53 [1.37 – 1.71] | <0.001 | 0.699  [0.658 – 0.739] |
| AVAD | 37 | 1.51 [1.32 – 1.73] | <0.001 | 0.620 [0.580 – 0.661] | 16 | 1.35 [1.17 – 1.56] | <0.001 |  |

Univariable and multivariable cox regression was used to evaluate AVAD and E/e’ as predictors of 5-year cardiovascular mortality. Wald’s chi-square values were used to compare the strength of association of different variables within regression models. The hazard ratio for E/e’ ratio is scaled by standard deviation increment, and AVAD by standard deviation decrement. The C-statistic of univariable and multivariable models was compared to evaluate differences in model discrimination. Abbreviations: AVAD = atrioventricular area difference; CI = confidence interval; E/e’ = E to septal e’ velocity ratio; HR = hazard ratio; LVEF = left ventricular ejection fraction.

### **Supplemental Table 3. Atrioventricular area difference and diastolic function grading as predictors of 5-year all-cause mortality in left ventricular ejection fraction subgroups**

| 75% ≤ LVEF n = 296, 82 events  5.0 [3.8–5.0] yrs follow up | **Univariable Model** | | | | **Multivariable Model** | | | |
| --- | --- | --- | --- | --- | --- | --- | --- | --- |
| **Variable** | Chi-Square | HR [95% CI] | p value | C-statistic  [95% CI] | Chi-Square | HR [95% CI] | p value | C-statistic  [95% CI] |
| Diastolic Dysfunction | 5 | 1.73 [1.07 – 2.80] | 0.03 | 0.572  [0.516 – 0.628] | 2 | 1.47 [0.88-2.47] | 0.14 | 0.608  [0.547 – 0.669] |
| Indeterminate Diastolic Function | 1 | 0.74 [0.39 – 1.44] | 0.38 |  | 1 | 0.68 [0.35-1.32] | 0.26 |  |
| AVAD | 6 | 1.32 [1.05 – 1.66] | 0.02 | 0.588  [0.526 – 0.649] | 4 | 1.26 [0.99-1.61] | 0.06 |  |
| 60% ≤ LVEF < 75% n = 3786, 840 events  5.0 [5.0-5.0] yrs follow up | **Univariable Model** | | | | **Multivariable Model** | | | |
| **Variable** | Chi-Square | HR [95% CI] | p value | C-statistic | Chi-Square | HR [95% CI] | p value | C-statistic |
| Diastolic Dysfunction | 235 | 3.25 [2.80 – 3.78] | <0.001 | 0.614  [0.597 – 0.631] | 168 | 2.86 [2.44 – 3.35] | <0.001 | 0.655  [0.637 – 0.673] |
| Indeterminate Diastolic Function | 20 | 1.58 [1.30 – 1.93] | <0.001 |  | 17 | 1.52 [1.25 – 1.86] | <0.001 |  |
| AVAD | 84 | 1.36 [1.27 – 1.45] | <0.001 | 0.595  [0.576 – 0.614] | 26 | 1.20 [1.12 – 1.29] | <0.001 |  |
| 50% ≤ EF < 60% n = 1094, 291 events  5.0 [4.6 – 5.0] yrs follow up | **Univariable Model** | | | | **Multivariable Model** | | | |
| **Variable** | Chi-Square | HR [95% CI] | p value | C-statistic | Chi-Square | HR [95% CI] | p value | C-statistic |
| Diastolic Dysfunction | 51 | 2.62 [2.01 – 3.41] | <0.001 | 0.595  [0.564 – 0.625] | 37 | 2.33 [1.77 – 3.05] | <0.001 | 0.632  [0.601 – 0.664] |
| Indeterminate Diastolic Function | 4 | 1.38 [1.01 – 1.88] | 0.04 |  | 2 | 1.28 [0.94 – 1.75] | 0.12 |  |
| AVAD | 29 | 1.30 [1.18 – 1.43] | <0.001 | 0.586  [0.555 – 0.617] | 16 | 1.23 [1.11 – 1.35] | <0.001 |  |

Survival analyses were repeated in LVEF subgroups of 50% ≤ LVEF < 60%, 60% ≤ LVEF < 75% and 75% ≤ LVEF to account for the influence of mitral annular plane systolic excursion on our measurement of AVAD. A similar trend in results were observed in all groups. Diastolic function grading was determined using the 2016 ASE/EACVI guidelines. Wald’s chi-square values were used to compare the strength of association of different variables within regression models. The hazard ratios for diastolic dysfunction and indeterminate diastolic function are reported with normal diastolic function as a reference level. The hazard ratio for AVAD is scaled by standard deviation decrement. The C-statistic of univariable and multivariable models was compared to evaluate differences in model discrimination. Abbreviations: ASE = American Society of Echocardiography; AVAD = atrioventricular area difference; CI = confidence interval; EACVI = European Association of Cardiovascular Imaging; HR = hazard ratio; LVEF = left ventricular ejection fraction.

### **Supplemental Table 4. Atrioventricular area difference and E/e’ as predictors of 5-year all-cause mortality in left ventricular ejection fraction subgroups**

| 75% ≤ LVEF n = 296, 82 events  5.0 [3.8–5.0] yrs follow up | **Univariable Model** | | | | **Multivariable Model** | | | |
| --- | --- | --- | --- | --- | --- | --- | --- | --- |
| **Variable** | Chi-Square | HR [95% CI] | p value | C-statistic  [95% CI] | Chi-Square | HR [95% CI] | p value | C-statistic  [95% CI] |
| E/e’ ratio | 10 | 1.35 [1.12 – 1.62] | 0.001 | 0.588  [0.527 – 0.650] | 6 | 1.28 [1.05 – 1.55] | 0.01 | 0.609  [0.548 – 0.670] |
| AVAD | 6 | 1.32 [1.05 – 1.66] | 0.02 | 0.588  [0.526 – 0.649] | 2 | 1.20 [0.94 – 1.53] | 0.14 |  |
| 60% ≤ LVEF < 75% n = 3786, 840 events  5.0 [5.0–5.0] yrs follow up | **Univariable Model** | | | | **Multivariable Model** | | | |
| **Variable** | Chi-Square | HR [95% CI] | p value | C-statistic | Chi-Square | HR [95% CI] | p value | C-statistic |
| E/e’ ratio | 249 | 1.57 [1.48 – 1.65] | <0.001 | 0.641  [0.623 – 0.659] | 176 | 1.49 [1.40 – 1.58] | <0.001 | 0.657  [0.639 – 0.675] |
| AVAD | 84 | 1.36 [1.27 – 1.45] | <0.001 | 0.595  [0.576 – 0.614] | 31 | 1.23 [1.14 – 1.31] | <0.001 |  |
| 50% ≤ EF < 60% n = 1094, 291 events  5.0 [4.6–5.0] yrs follow up | **Univariable Model** | | | | **Multivariable Model** | | | |
| **Variable** | Chi-Square | HR [95% CI] | p value | C-statistic | Chi-Square | HR [95% CI] | p value | C-statistic |
| E/e’ ratio | 69 | 1.39 [1.28 – 1.50] | <0.001 | 0.633  [0.602 – 0.663] | 47 | 1.33 [1.22 – 1.44] | <0.001 | 0.652  [0.622 – 0.682] |
| AVAD | 29 | 1.30 [1.18 – 1.43] | <0.001 | 0.586  [0.555 – 0.617] | 12 | 1.21 [1.09 – 1.34] | <0.001 |  |

Survival analyses were repeated in LVEF subgroups of 50% ≤ LVEF < 60%, 60% ≤ LVEF < 75% and 75% ≤ LVEF to account for the influence of mitral annular plane systolic excursion on our measurement of AVAD. A similar trend in results were observed in all groups. Wald’s chi-square values were used to compare the strength of association of different variables within regression models. The hazard ratio for E/e’ ratio is scaled by standard deviation increment, and AVAD by standard deviation decrement. The C-statistic of univariable and multivariable models was compared to evaluate differences in model discrimination. Abbreviations: AVAD = atrioventricular area difference; CI = confidence interval; C-statistic = concordance statistic; E/e’ = E to septal e’ velocity ratio; HR = hazard ratio; LVEF = left ventricular ejection fraction.

### **Supplemental Table 5. Univariable association between left ventricular ejection fraction and atrioventricular area difference in left ventricular ejection fraction subgroups**

| **LVEF group** | **R^2^** | **p value** |
| --- | --- | --- |
| LVEF ≥ 75% | 0.01 | 0.03 |
| 60% ≤ LVEF < 75% | 0.01 | <0.001 |
| 50% ≤ LVEF < 60% | 0.01 | 0.001 |
| LVEF ≤ 50% | 0.03 | <0.001 |

Univariable linear regression was used to determine the association between AVAD and LVEF. Patients with a similar LVEF are known to have a comparable mitral annular plane systolic excursion, meaning the movement of the mitral annular plane does not meaningfully affect differences in AVAD between patients with a similar LVEF. Abbreviations: AVAD = atrioventricular area difference; LVEF = left ventricular ejection fraction.
